# Costochondritis syndrome and thoracic-chest related pain: a scoping review

**DOI:** 10.1101/2024.02.11.24302642

**Authors:** Andrea Bolandrini, Antonio Barile, Marco Segat, Martina Zaninetti, Matteo Fascia, Andrea Segat, Marta Sebastiani, Federico Minetti, Filippo Maselli, Giovanni Galeoto, Michele Margelli

## Abstract

**Background:** Costochondritis is a frequent diagnosis related to chest pain but the etiology, treatment, and evolution of the disease are poorly documented.

Clinical examination lacks specificity and most treatment recommendations are conservative in nature and have been traditionally accepted, perhaps because of the self-limited nature of the condition.

However, costochondritis that does not self-resolve is referred to as atypical costochondritis and is associated with high medical expenses and psychological burden on the patient.This scoping review will aim to provide a clear overview about costochondritis, including definitions and terms, signs and symptoms, causes, diagnosis, treatment and prognosis. This work will also aim to propose shareable terms to describe this condition.

**Inclusion Criteria:** Every study describing a condition of non-cardiac musculoskeletal chest pain due to inflammation of the costochondral junctions of ribs or costosternal joints where the areas of tenderness are not generally accompanied by heat, erythema or localized swelling (e.g.,Tietze syndrome). This scoping review will consider studies conducted in any context. Articles in English or Italian will be considered.

**Methods:** The proposed scoping review will be conducted in accordance with the Joanna Briggs Institute methodology (JBI) for scoping reviews.

The search will be carried out on 5 databases: MEDLINE, Embase, Cochrane Library, CENTRAL and CINAHL. Addictionally, research protocols will be searched on PROSPERO and ClinicalTrials.gov. Further research of grey literature will be carried out through OpenGrey, Open Science Framework and Grey Literature Network Service.

Selection and data extraction will be conducted by two blind independent researchers and inconsistencies will be resolved by a third reviewer.

The results will be presented in a schematic, tabular and descriptive format that will line up with the objectives and scope of the review.

**Conclusions:** This scoping review will aim to provide a comprehensive overview of the topic. The results will add meaningful information for clinicians. Furthermore, any knowledge gaps of the topic will be identified.

## BACKGROUND

Costochondritis, also known as costosternal syndrome, parasternal chondrodynia, or anterior chest wall syndrome (1), is a self-limiting, poorly described and benign condition that usually manifests as non-cardiac chest pain (2). It is a common condition seen in patients presenting to physician’s office or emergency department (3–6). Even though costochondritis is a frequent diagnosis related to chest pain, the etiology, treatment, and evolution of the disease are often poorly documented (3).

The typical presentation is parasternal pain; the upper (predominantly second through fifth) costosternal and/or costochondral junctions are most commonly involved; the areas of tenderness are not generally accompanied by heat, erythema or localized swelling (1,3–5,7). The primary symptom of costochondritis is chest wall pain of varying intensity, typically described as sharp, aching, or pressure-like. The pain is often exacerbated by upper body movement, deep breathing, and exertional activities (5).

The diagnosis is made based on the exclusion of other causes of chest pain and the ability to reproduce the pain by palpation to the area (4). However, in a study of costochondritis in an emergency setting 6% of patients with pain reproduced by chest wall palpation were also diagnosed with myocardial infarction (3). Because the clinical examination lacks specificity, most patients with chest pain still need a thorough evaluation, including electrocardiography at a minimum, to rule out other more serious causes (1,8–19).

There are no laboratory tests, imaging tests, or electrocardiography findings specifically for the diagnosis of costochondritis (20).

No high-quality studies have examined the effectiveness of any treatment options for costochondritis. Most treatment recommendations are conservative in nature and have been traditionally accepted, perhaps because of the self-limited nature of the condition (1,21). However, costochondritis that does not self-resolve is referred to as atypical costochondritis and is associated with high medical expenses and psychological burden on the patient (2).

This scoping review will aim to provide a clear overview about costochondritis, including definitions and terms, signs and symptoms, causes, diagnosis, treatment and prognosis. This work will also aim to propose shareable terms to describe this condition.

### Review question

What is the current state of the literature about costochondritis?

The main objectives of this study will be:

1. To identify the available definitions and terms used to describe costochondritis;
2. To propose shareable terms to describe this condition;
3. To map and describe the available literature regarding signs and symptoms, causes, diagnosis, treatment and prognosis;
4. To identify any possible knowledge gaps on these topics.

## METHODS

### Study design and protocol

The proposed scoping review will be conducted in accordance with the Joanna Briggs Institute methodology (JBI) for scoping reviews.

The Preferred Reporting Items for Systematic reviews and Meta-Analyses extension for Scoping Reviews (PRISMA-ScR) Checklist for reporting will be used, and it is priori registered at Medxriv (https://www.medrxiv.org).

### Search strategy

As recommended in all JBI types of reviews and PRISMA-S, a three-step search strategy is to be utilized.

1. The first step is an initial limited search of an appropriate online databases relevant to the topic (MEDLINE). This initial search is then followed by an analysis of the text words contained in the title and abstract of retrieved papers, and of the index terms used to describe the articles.
2. The second step concerns a search using all identified keywords and index terms should then be undertaken across all included databases.
3. The third step, the reference list of identified reports and articles should be searched for additional sources.

The search strategies were peer-reviewed by an experienced librarian and were further refined through team discussion. No search limitations and filters applied (language and time). Reviewers’ intent to contact authors of primary sources or reviews for further information. Complete search strategy for Medline is included as an appendix 1 to the protocol. Search strategy will be adapted to be used in other databases.

### Inclusion criteria

We will follow the acronym PCC to describe elements of the inclusion criteria:

— *Population* This scoping review will consider studies on living adult humans (aged 18 or above) with non-cardiac musculoskeletal chest pain.
— *Concept* This scoping review will consider research studies that describe a condition of non-cardiac musculoskeletal chest pain due to inflammation of the costochondral junctions of ribs or costosternal joints where the areas of tenderness are not generally accompanied by heat, erythema or localized swelling (e.g., Tietze syndrome).
— *Context* This scoping review will consider studies conducted in any context.

#### Sources

This scoping review will consider any study designs or publication type for inclusion. No date and geographical limits will be used. We will consider articles in English and Italian. Studies that do not meet the above-stated Population-Concept-Context (PCC) criteria or which provide insufficient information will be excluded.

#### Study selection

For the selection process, the first thing to do is to select a sample of 25 title/abstracts that will be analyzed from the entire team using eligibility criteria and definitions/elaboration document. Then, the team will discuss discrepancies and make modifications to the eligibility criteria and definitions/elaboration document. The team will only start screening when 75% (or greater) of agreement between reviewers will be achieved. To start the screening, we will use GoogleSheets.

The selection phase will begin assessing all the titles and abstracts of the studies retrieved using the search strategy and those from additional sources. These studies will be screened independently by two review authors, to identify those that potentially meet the inclusion criteria. Any disagreement will be solved by two reviewer consensus. Subsequently, the full text of these potentially eligible studies will be independently assessed for eligibility by the two reviewers. The reasons for excluding articles will be recorded. We developed a google form (charting table) containing the elements to standardize the selection.

There should be a narrative description of the selection process accompanied by a flowchart of review process (from the PRISMA-ScR statement).

## Data extraction

Extraction module will be reviewed, before the implementation, by the research team and pre-tested to ensure that the form accurately captures the information. Modifications will be detailed in the full scoping review.

Data extraction will be conducted by two blind independent researchers and inconsistencies will be resolved by a third reviewer.

### Data items

Key information will be described in a charting table with the description of: author; publication year; country of conduct of the study; setting of conduct of the study; methodology/type of study, aims of the study, population characteristics from which patients are extracted (including gender, age and social variables); Concept: synonyms of costochondritis; definition of costochondritis, signs and symptoms, diagnostics, treatment strategies, prognostics.

### Critical appraisal of individual sources of evidence

Not provided.

### Data management

As a scoping review, the purpose is to aggregate the findings and to present an overview of the research rather than to evaluate the quality of the individual studies. The results will be presented as a map of data, which are extracted from different documents. These will be included in a schematic, tabular and descriptive format that will line up with the objectives and scope of the review. Descriptive analysis will consist of a distribution of the evidence sources by year or period of publication, country of origin, area of intervention (clinical, political, educational, etc.) and research methods. Results will be presented such as: definitions, evaluation and treatment.

### Data Availability

Due to the nature of this research, participants of this study did not agree for their data to be shared publicly, so supporting data is not available.

## Appendix 1 Complete search strategy for Medline

Search: “costochondritis” OR “costosternal syndrome” OR “costosternal chondrodynia” OR “parasternal chondrodynia” OR “tietze syndrome” OR “tietze’s syndrome” OR “costal chondritis” OR “chest wall syndrome”

